# Etuvetidigene autotemcel for the treatment of Wiskott-Aldrich Syndrome

**DOI:** 10.64898/2025.11.25.25340584

**Authors:** Francesca Ferrua, Sabina Cenciarelli, Stefania Giannelli, Stefania Galimberti, Shanmuganathan Chandrakasan, Federico Fraschetta, Carmen Caputo, Davide Sala, Ilaria Monti, Federica Barzaghi, Valeria Calbi, Daniele Canarutto, Giulia Consiglieri, Matteo Doglio, Francesca Fumagalli, Vera Gallo, Maddalena Migliavacca, Salvatore Recupero, Francesca Tucci, Alessia Orsini, Raffaella Milani, Mariam Datukishvili, Simona De Gregori, Eugenio Montini, Paolo Silvani, Matias Soncini, Elena Tomasetto, Koen van Rossem, Laura Castagnaro, Federica Miotto, Stefano Zancan, Celeste Scotti, Sean Russell, Luigi Naldini, Fabio Ciceri, Maria Ester Bernardo, Suhag Parikh, Maria Pia Cicalese, Alessandro Aiuti, the WAS gene therapy group

**Affiliations:** San Raffaele Telethon Institute for Gene Therapy (SR-Tiget), IRCCS San Raffaele Scientific Institute, Milan, Italy; Pediatric Immunohematology Unit and Bone Marrow Transplantation Unit, IRCCS San Raffaele Scientific Institute, Milan, Italy; Department of Biomedicine and Prevention, PhD in Immunology, Molecular Medicine and Applied Biotechnology, University of Rome Tor Vergata, Rome, Italy; Bicocca Bioinformatics Biostatistics and Bioimaging B4 Center, School of Medicine and Surgery, University of Milano - Bicocca, Monza, Italy; Biostatistics and Clinical Epidemiology, Fondazione IRCCS San Gerardo dei Tintori, Monza; Aflac Cancer and Blood Disorders Center, Children’s Healthcare of Atlanta, Emory University School of Medicine, Atlanta, USA; Vita-Salute San Raffaele University, Milan, Italy; Neurology Unit and Neurophysiology Service, IRCCS San Raffaele Scientific Institute, Milan, Italy; Immunohematology and Transfusion Medicine Unit, IRCCS San Raffaele Scientific Institute, Milan, Italy; Department of diagnostic medicine: clinical chemical analysis laboratory; Fondazione IRCCS Policlinico San Matteo, Pavia, Italy; Department of Anesthesia and Critical Care, IRCCS San Raffaele Scientific Institute, Milan, Italy; Fondazione Telethon ETS, Rome, Italy; Karos Healthcare Consulting, Vosselaar, Belgium; PrimeRA Pharma Partners, Nottingham, UK; Hematology and Bone Marrow Transplantation Unit, IRCCS San Raffaele Scientific Institute, Milan, Italy

**Keywords:** gene therapy, lentiviral vector, hematopoietic stem cell, immune deficiency, platelet, transplantation

## Abstract

**BACKGROUND:** Wiskott-Aldrich Syndrome (WAS) is a rare, X-linked, life-threatening inborn error of immunity and platelet disorder caused by WAS protein (WASP)-encoding gene mutations. Etuvetidigene autotemcel (etu-cel) is an autologous gene therapy (GT) consisting of hematopoietic stem progenitor cell (HSPCs) transduced *ex vivo* with a lentiviral vector encoding human WAS cDNA.

**METHODS:** Etu-cel was intravenously infused after rituximab and reduced-intensity conditioning. Data from WAS patients treated in two prospective open-label clinical trials (phase I/II n=8; phase III n=10) and one expanded access program (EAP) (n=9) were integrated to evaluate efficacy and safety of etu-cel. Primary efficacy endpoints were overall survival, rate of severe infections from 6 to 18 months after GT and rate of moderate/severe bleeding episodes in the first 12 months post-treatment compared with 1 year prior to GT. Secondary efficacy endpoints included engraftment of gene-corrected cells, WASP expression, T-cell function, platelet count, autoimmunity and eczema over time. Safety endpoints included adverse events (AEs), immune response to transgene, development of replication-competent lentivirus (RCL) and abnormal clonal proliferation (ACP).

**RESULTS:** Median follow-up was 5.7 years (range: 0.4-13.3). Median age at treatment was 2.6 years (range: 1.0-35.1). Overall survival was 96%; one EAP subject died early post-GT due to deterioration of a pre-existing neurological condition. The rate of severe infections per person-year of observation (PYO) decreased from 2.00 (95% CI: 1.50-2.61) pre-GT to 0.15 (95% CI: 0.04-0.39) in the 6-18 months period post-GT. The rate of moderate and severe bleeding events per PYO decreased from 2.00 (95% CI: 1.50-2.61) to 0.80 (95% CI: 0.49-1.22) in the 12 months after GT. After GT, multilineage engraftment of gene-corrected cells was sustained over time. WASP expression in platelets and lymphocytes increased. Platelet count, T-cell functionality, eczema and autoimmunity improved. The most common adverse event ≥ grade 3 was device related infection. Etu-cel was well-tolerated with no treatment-related adverse events and no evidence of insertional oncogenesis.

**CONCLUSIONS:** With up to 13 years follow-up, etu-cel demonstrates a favorable benefit-risk profile, leading to sustained long-term clinical benefit.

(Funded by GlaxoSmithKline [GSK], Orchard Therapeutics, Fondazione Telethon; ClinicalTrials.gov numbers: NCT01515462, NCT03837483)

## INTRODUCTION

Wiskott-Aldrich Syndrome (WAS) is a rare, X-linked, life-threatening inborn error of immunity and platelet disorder caused by WAS protein (WASP)-encoding gene mutations^1–3^. WAS is characterized by thrombocytopenia, bleeding events, recurrent and severe infections, eczema, and increased risk of immune dysregulation and malignancy^4^, leading to significantly reduced life expectancy. Genotype can be a predictive biomarker for phenotype severity, including patients’ survival and severe events presentation^5^. Current treatment options consist of symptomatic and preventive management and allogeneic hematopoietic stem cell transplantation (HSCT). Supportive treatments include bleeding prophylaxis with platelet transfusions, use of antifibrinolytic agents or off-label thrombopoietin (TPO) agonists, infection prophylaxis and immunoglobulin (Ig) replacement therapy (IgRT), and early treatment with antimicrobials. Autoimmune and autoinflammatory manifestations are managed with immunosuppressive drugs, corticosteroids, rituximab, and *off label* use of anti-IL1 agents^4,6^.

HSCT in conjunction with myeloablation and immune suppression can be curative, leading to restoration of WASP expression, platelet count, immune functions, and resolution of eczema with 3 to 5-year survival rates ∼90%^7,8^. However, despite improvement in recent years, HSCT is still hampered by limited donor availability and potential complications, including infections, autoimmune manifestations, graft failure or graft rejection, and acute or chronic graft-versus-host disease (GvHD)^7,8^. In addition, HSCT from patients transplanted at ≥5 years of age or from mismatched family donors (MMFD) is still associated with substantial morbidity and increased mortality^7,8^.

Etuvetidigene autotemcel (etu-cel) is an autologous gene therapy (GT) consisting of hematopoietic stem and progenitor cells (HSPCs) transduced *ex vivo* with a lentiviral vector encoding human WAS cDNA under the control of a reconstituted promoter from the endogenous locus^9^. We previously reported interim safety and efficacy findings in 8 WAS subjects treated in a phase I/II clinical trial with a median follow-up of 3.6 years^10^. We now report the results of an integrated efficacy and safety analysis in 27 WAS patients treated with etu-cel with extended long-term follow-up.

## METHODS

### PATIENT POPULATION AND STUDY CONDUCT

We integrated data from 2 prospective open-label clinical trials (a phase I/II study TIGET-WAS [n=8, EUDRACT 2009-017346-32, NCT01515462] and a two-center phase III study OTL-103-4 [n=10, EUDRACT 2018-003842-18, NCT03837483]), and from an expanded access program (EAP) (n=10). Of the 28 enrolled WAS patients, 27 were treated with etu-cel (Table 1, Table S1 and Fig. S1). One patient of the EAP did not receive conditioning or etu-cel because the target number of CD34+ cells for drug product (DP) manufacturing was not reached (Fig. S1).

**Table 1.** Summary of Patient and Drug Product Characteristics in the Efficacy Population.

| Characteristics | Etu-cel (n=27) |
| --- | --- |
| Age at disease onset in months, median (range) | 0.3 (0.0, 32.3) |
| Age on date of gene therapy in years, median (range) | 2.6 (1.0, 35.1) |
| Age group (5 year cut-off categories) on date of gene therapy, n (%) |  |
| - <5 years | 18 (66.7) |
| - ≥5 years | 9 (33.3) |
| Zhu score at baseline, n (%) |  |
| - 2.0 | 1 (3.7) |
| - 3.0 | 13 (48.1) |
| - 4.0 | 4 (14.8) |
| - 5.0A | 8 (29.6) |
| - Missing* | 1 (3.7) |
| WAS gene mutation at baseline, n (%) |  |
| - Class I | 5 (18.5) |
| - Class II | 22 (81.5) |
| Duration of follow-up in years, median (range)<sup>§</sup> | 5.7 (0.4, 13.3) |
| Duration of follow-up in years in patients alive at the data cut-off date, median (range) | 5.7 (2.3, 13.3) |
| Transduction efficiency in the DP, n (%) |  |
| - ≤80 % | 6 (22.2) |
| - >80 % | 21 (77.8) |
| Vector Copy Number (copies per cell) in the DP, median (range) | 2.3 (0.9 – 4.3) |
| Number of CD34+ cells infused (10<sup>6</sup>/kg), median (range) | 16.9 (7.0 – 31.0) |

WAS denotes Wiskott-Aldrich Syndrome; n = number of subjects
\* A baseline Zhu score of 5.0A in one participant was windowed to the treatment visit because it was assessed after the start of mobilization for peripheral blood stem cell collection
§ This includes the available follow-up of one adult patient who died 4.5 months after GT due to a serious adverse event (SAE) of neurological decompensation, that was considered unrelated to etu-cel.

Patients received fresh (n=17) or cryopreserved (n=10) formulation of etu-cel. The first patient was treated on 11 June 2010 and the last patient was treated on 26 Sep 2022; data were collected between Apr 2010 and Jan 2025. The patient sample contained patients from across the world, with different genotypes, ethnic and racial backgrounds (Table S2) and is representative of the broader population of WAS patients. Patients were treated and followed at Ospedale San Raffaele (OSR), in Milan, Italy (n=26) and Children’s Healthcare of Atlanta (CHOA), Atlanta, United States (n=1). The clinical trials and EAP included similar methods and schedules of assessments, allowing for an integrated analysis. Studies conducted at OSR were approved by the OSR Ethics Committee and the Agenzia Italiana del Farmaco; study performed at CHOA was approved by local Institutional Review Board (IRB) and conducted under an active investigational new drug (IND) with US Food and Drug Administration. The studies were undertaken in a manner consistent with the principles of Good Clinical Practice and the Declaration of Helsinki. Written informed consent was obtained from patients, parents, or guardians.

Key eligibility criteria included patients with diagnosis of WAS defined by genetic mutation with no HLA-identical related donor available and at least one of the following criteria: severe WAS gene mutation, absent WASP expression, and/or severe clinical score (Zhu clinical score ≥ 3)^11^. The detailed inclusion and exclusion criteria of the specific studies are reported in Table S3.

### PROCEDURES

WAS patients were treated with etu-cel manufactured starting from autologous CD34+ HSPC purified from either bone marrow (BM) harvest (n=5), mobilized peripheral blood (mPB) apheresis (n=21) following granulocyte colony-stimulating factor with (n=20) or without (n=1) plerixafor administration, or from both sources (BM+mPB, n=1) (see Supplementary Methods). CD34+ cells were transduced *ex vivo* with a self-inactivating lentiviral vector (LV) LV-w1.6WAS encoding WAS cDNA under the control of a reconstituted promoter from the endogenous locus, as previously described^9,10^. Participants in study TIGET-WAS and patients in the EAP received the fresh formulation of etu-cel, whereas participants in study OTL-103-4 received the cryopreserved formulation. Etu-cel was intravenously infused after rituximab and reduced-intensity conditioning (RIC) with weight-based and pharmacokinetic adjusted-busulfan and fludarabine (Fig. S2, Table S4).

After treatment, patients were followed up by clinical, laboratory and instrumental assessments, as defined by the respective clinical study or program.

### ENDPOINTS AND STATISTICAL ANALYSES

Primary efficacy endpoints were overall survival (OS), rate of severe infections from 6 to 18 months post-GT and rate of moderate/severe bleeding events in the first 12 months post-GT compared with 1 year prior to GT. Secondary efficacy endpoints included sustained engraftment of gene-corrected cells, WASP expression in lymphocytes and platelets, T-cell function, platelet count, autoimmunity and eczema over time. Safety endpoints included adverse events (AEs), immune response to transgene, development of replication-competent lentivirus (RCL) and abnormal clonal proliferation (ACP).

Statistical analyses are described in detail in the Supplementary Methods. The Integrated Summary of Safety (ISS) comprised 28 enrolled patients including one patient who did not receive treatment with etu-cel. The Integrated Summary of Efficacy (ISE) included all 27 patients treated with etu-cel (Fig. S1). Due to the ultra-rare nature of the disease, the sample sizes for the studies were determined on practical and feasibility considerations rather than statistical arguments. All analyses were based on descriptive statistics for all the endpoints (safety and efficacy); no formal hypothesis testing was performed, but, when applicable, 95% Confidence Intervals (CI) were calculated to quantify statistical uncertainty in the estimates.

## RESULTS

### PATIENTS

Demographic features and DP characteristics of 27 WAS patients who were treated with etu-cel are described in Table 1 and Table S2. Median age at treatment was 2.6 years (range: 1.0-35.1) with 9 out of 27 patients being ≥5 years of age. All patients except one had a severe disease score and most (81.5%) had a severe (class II) genotype. The median cell dose was 16.9×10^6/kg CD34+ cells (range: 7-31x10^6/kg), with a median vector copy number (VCN) per cell of 2.3 (range: 0.9-4.3) (Table 1).

### PRIMARY ENDPOINTS

Overall survival (OS) was 96% at 1 and 5 years of follow-up (95%CI: 76-99%) (Fig. 1A); one EAP subject died 4.5 months post-GT due to deterioration of a pre-existing neurological condition. Median follow-up in surviving patients was 5.72 years (range: 2.31-13.26 years).The rate of severe infections per person-year of observation (PYO) decreased from 2.00 (95%CI: 1.50-2.61) in the 12 months before GT to 0.15 (95%CI: 0.04-0.39) in the 6-18 months post-GT and 0.05 (95%CI: 0.01-0.16) in the >5-year post-treatment period (Fig. 1B, Table S5). The most frequently reported severe infections were device-related. Of note, Cytomegalovirus (CMV) reactivation requiring antiviral treatment was no longer observed after 6 months post-GT (Table S6).

**Figure 1.**
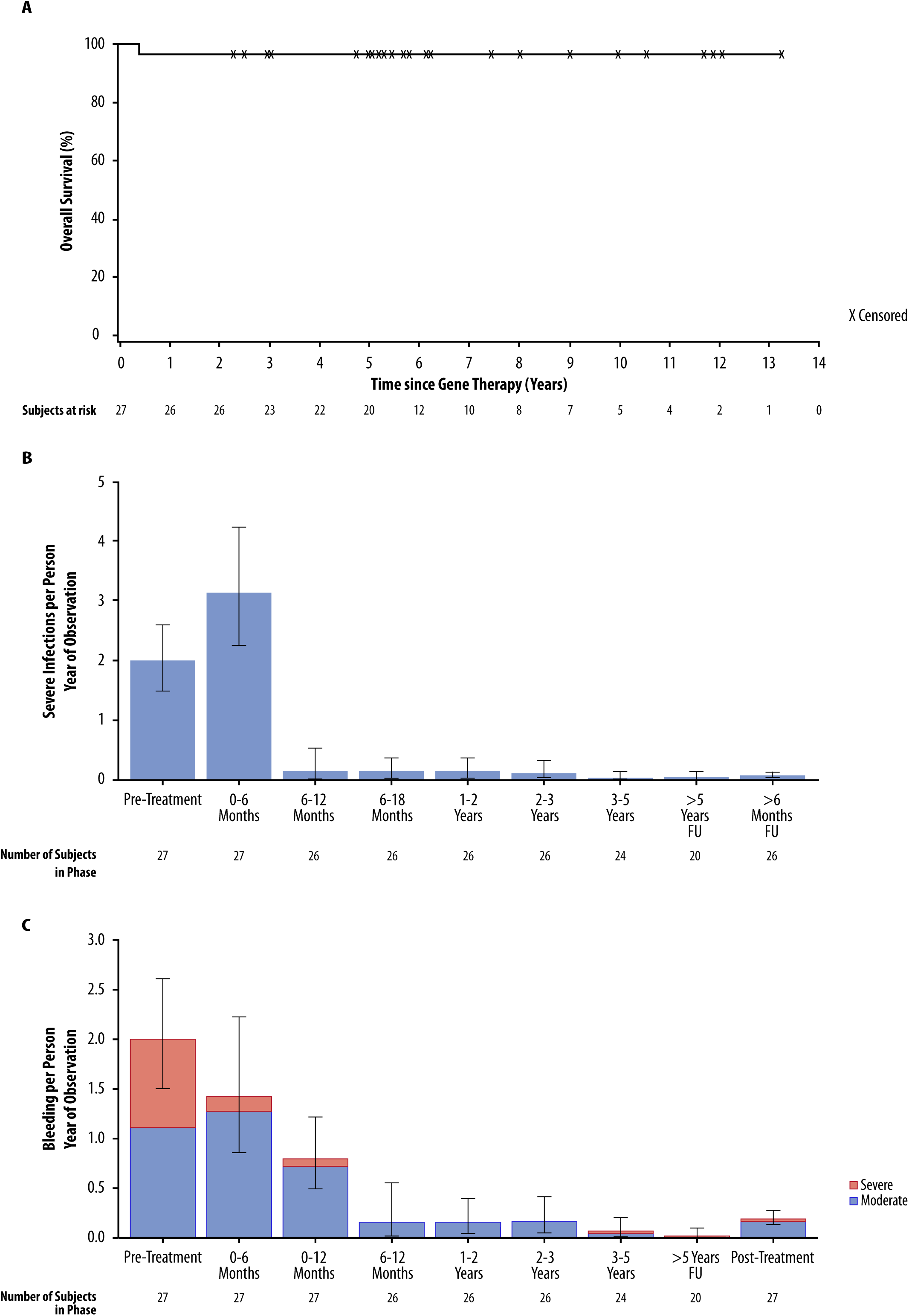
Primary efficacy endpoints (Efficacy population) **(A) Kaplan-Meier plot of overall survival. Participants are censored at the latest timepoint for which vital status information is available (up to the data cut-off).** **(B) Rate of severe infections by treatment phase.** The rate of events was estimated as the number of events over person-years of observation. Error bars indicate the 95% CI. Events on the day of etu-cel infusion are not included. **(C) Rate of moderate and severe bleeding events by treatment phase.** Confidence interval for bleeding is for combined moderate and severe rate. The rate of events was estimated as the number of events over person-years of observation. Error bars indicate the 95% CI. Events on the day of etu-cel infusion are not included. Note: FU=follow-up. Pre-Treatment: events occurring in the 12 months before GT. At >6 months FU: events occurring more than 6 months after etu-cel infusion to last follow-up. At >5 years FU: events occurring more than 5 years after etu-cel infusion to last follow-up. Post-treatment: events occurring after gene therapy until latest follow-up (up to 13.3 years).

The rate of moderate and severe bleeding events per PYO decreased from 2.00 (95%CI: 1.50-2.61) in the 12 months before GT to 0.80 (95%CI: 0.49-1.22) in the 12 months after GT and 0.02 (95%CI: 0.00−0.10) in the >5-year post-treatment period (Fig. 1C, Table S7).

The efficacy of etu-cel in protecting patients from severe infections and moderate/severe bleedings was confirmed at individual patient level across different studies and maintained over time up to the latest available follow-up (Fig. S3).

### ENGRAFTMENT OF GENE CORRECTED CELLS AND WASP EXPRESSION

All participants experienced transient grade 4 neutropenia (absolute neutrophil counts <500/μL) after RIC, with neutrophil engraftment achieved at a median of 23 days post-GT (Fig.S4). There were no engraftment failures and no patients required reinfusion of untransduced autologous cell back up. All participants achieved adequate engraftment of LV transduced BM CD34^+^ and/or PB CD3^+^ cells (Supplementary Methods, Fig.S5). Multilineage engraftment in PB and BM was stable up to the latest follow-up (Fig. 2A, Fig. S6A). The percentage of LV+ clonogenic progenitors in the BM showed evidence of robust engraftment of transduced progenitors, remaining stable throughout follow-up (Fig. S6B, Table S8).

**Figure 2.**
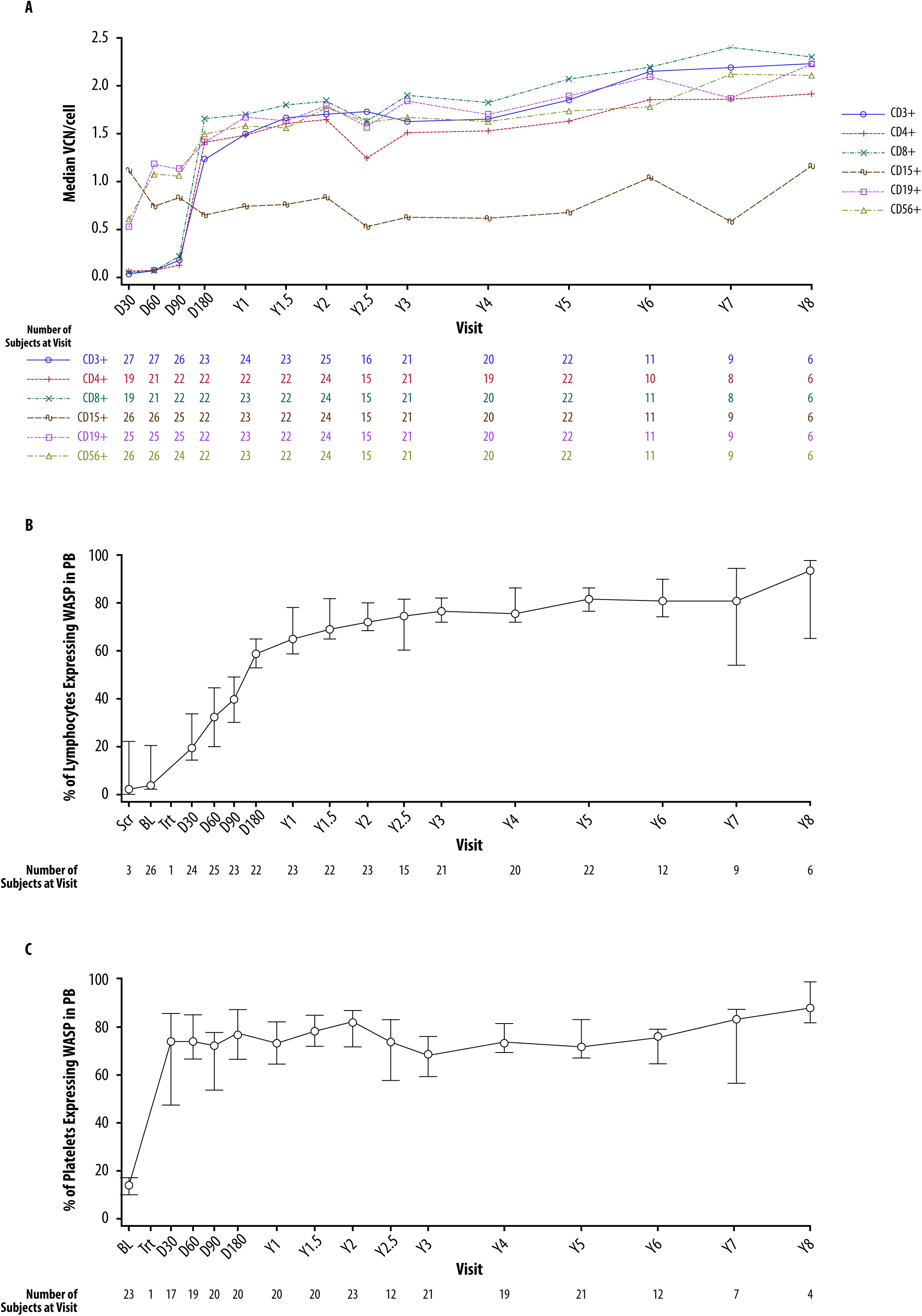
Engraftment of gene-corrected cells and WASP expression restoration after treatment with etu-cel (Efficacy population) **(A) Median values for VCN/cell in peripheral blood cell lineages.** D=Day; CD=cluster of differentiation; VCN=vector copy number; Y=Year. Results based on fewer than two participants were omitted. **(B-C) Median (95% CI) percentages of lymphocytes (B) and platelets (C) in PB expressing WASP assessed by flow cytometry over time.** Results based on fewer than two participants were omitted. Note: PB= peripheral blood, Scr= screening, BL=baseline, Trt=Treatment, D= Day, Y=Year, WASP= Wiskott-Aldrich Syndrome Protein. Transduced cells infusion (gene therapy) was performed at the Treatment Visit (Trt).

An exploratory analysis of covariates for efficacy showed a positive correlation between DP VCN/cell and engraftment of gene corrected cells at 1 year (Fig. S7).

The engraftment of gene corrected cells led to restoration of WASP expression in lymphocytes (including T, B and NK cells), platelets and monocytes, which increased mostly in the first 6 months then progressively reached a plateau and remained stable over time (Fig. 2B, 2C, and Fig. S8). Median VCN/cell and WASP expression were higher in the lymphoid than the myeloid lineage. This could be expected, as submyeloablative conditioning leads to only partial reconstitution of the myeloid compartment, whereas the selective advantage of WASP-expressing lymphoid cells accounts for a more extended reconstitution of lymphoid lineages with a predominance of corrected cells (Fig. 2, Fig. S8, Table S9).

### IMMUNE RECONSTITUTION

Before GT, most patients displayed lymphopenia, mainly involving B cells, CD8+ T cells and naive CD4+ T cells. Thanks to immune reconstitution of gene corrected cells, normalization of these subpopulations was observed in the majority of patients starting from 1 year follow-up (Table S10). All participants had reduced *in vitro* lymphocyte proliferative responses to CD3 stimulation at baseline which increased over time, reaching a median response within normal range at anti-CD3i concentration of 1 μg/mL from year 1 (Fig. 3A). Sustained immunoglobulin replacement therapy could be stopped in all patients who were alive at latest follow-up (26 out of 27 treated with etu-cel), with a median time to cessation of 320.5 days (range: 91–1843 days) (Fig. 3B). Cessation of sustained antimicrobial treatment was achieved in 25 (92.6%) patients, with a median time to cessation of 405 days (range: 138-2534 days) (Supplementary Fig. S9). All 26 patients treated with etu-cel and alive started a vaccination schedule post-GT after at least 3 months since last immunoglobulin administration. The majority of them were able to mount a protective response to most of the vaccinal antigens tested, including live attenuated vaccines (Table S11). In addition, 3 patients experienced VZV or measles infection before receiving the specific vaccine and demonstrated an adequate specific antibody response to the native virus, without complications.

**Figure 3.**
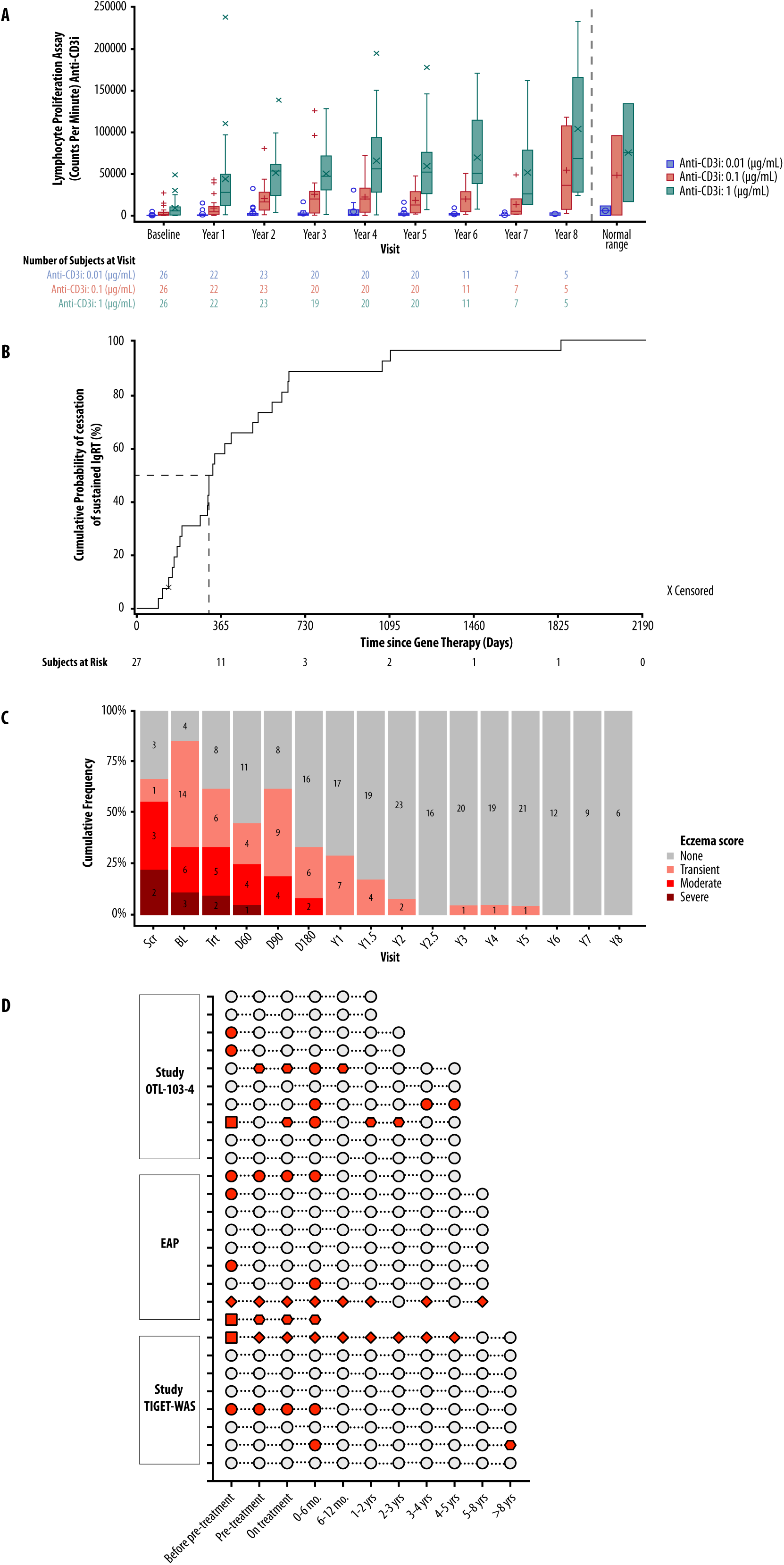
Immune reconstitution after treatment with etu-cel (Efficacy population) **(A) Boxplot of T-cell proliferation responses to CD3 stimulation at baseline and each year after etu-cel infusion.** Each box indicates the interquartile range, with the lower end of the box being the first quartile (Q1) and the upper end is the third quartile (Q3). The T-shaped whiskers represent 1.5 times the interquartile range. Large symbols (o, +, x) represent the mean, whereas small symbols represent outliers. Horizontal line within box represents the median. The three different colors represent the three concentrations of anti-CD3i monoclonal antibody (mAb) used to test dose-response of T-cell proliferation *in vitro*. Note: CD3i=immobilized CD3. **(B) Kaplan-Meier plot of time to cessation of sustained immunoglobulin replacement therapy (Efficacy population).** The dashed line represents the median time of cessation of sustained immunoglobulin replacement therapy (IgRT). **(C) Percentage stacked barchart of eczema score over time.** This graph represents the cumulative frequency (%) of subjects having different eczema scores at various timepoints before and after treatment with etu-cel. The absolute number of subjects having a certain score at a given timepoint is reported in the relative portion of the column relative to that timepoint. Only subjects for which eczema score was available are represented in the graph. Eczema Scores: 1=None, 2=Transient, 3=Moderate, 4=Severe. Note: transduced cells infusion (gene therapy) was performed at the Treatment Visit (Trt). Scr = Screening, BL = Baseline, Trt = Treatment, D = Day, Y = Year. **(D)** Autoimmune events prior and after etu-cel infusion (Efficacy population) This graph represents the presence of absence of autoimmune clinical manifestations over time, before and after etu-cel infusion. Each row represents a patient, enrolled in either the TIGET-WAS or OTL-103-4 clinical studies or in the EAP. Grey dots represent absence of autoimmune manifestation in the indicated time window. Red symbols show the presence of one or more autoimmune (AI) manifestations in the indicated time window: a round dot represents AI cytopenia, rhombus represents inflammatory bowel disease, hexagon indicates other types of clinical autoimmunity (including Henoch-Schonlein purpura, IgA nephropathy, psoriasis, uveitis), square indicates the simultaneous presence of multiple autoimmune manifestations. Additional details about autoimmune events before and after etu-cel infusion are described in Table S13 and S14. Pre-treatment indicates the period from the date of screening up to and including the day before the on-treatment phase. On-treatment denotes the period from Day -22 or the day of PBSC mobilization, if performed, up to and including Day 1 (day of gene therapy).

After GT, social life improved progressively: from 3 years after GT, all participants had stopped living in a protected environment, >80% attended school/kindergarden and about half of them were practicing sport. After GT, the annualized rate of hospitalizations, decreased from 2.85 per PYO in the 12 months before GT to 0.46 per PYO in the 6-12--month period post-GT, and decreased further to 0.09 hospitalizations per PYO in the >5-year period (Table S12).

### IMMUNE DYSREGULATION

Twenty-three out of 27 patients had eczema at baseline. No participant had an eczema score^12^ of either “moderate” or “severe” from year 1 onwards. At year 2, 23 (92.0%) participants were free of eczema and from year 6 onward, no participants had eczema (Fig. 3C).

Ten patients experienced clinical manifestations of autoimmunity in their clinical history before GT. Clinical manifestations of autoimmunity ongoing at treatment completely resolved after etu-cel infusion, except for 1 patient, who experienced a reactivation of ulcerative colitis at 3.6 and 7.3 years after GT. After treatment, 5 subjects (18.5%) experienced transient manifestations of autoimmune cytopenia in the first 6 months of FU, which then resolved. In the subsequent period (>6 months post-GT) autoimmune manifestations were observed in 4 patients (14.8%) which required short-term or no intervention. (Fig. 3D, Table S13 and S14).

Nine patients experienced autoinflammatory clinical manifestations before GT. One patient continued to experience autoinflammatory manifestations after GT, subsequently diagnosed with concomitant Familial Mediterranean Fever. Three patients received anti-IL1 agents to control the manifestations before GT^6,13^ and no relapse was observed after etu-cel infusion, despite discontinuation of anti-IL1 agents. After GT, transient *de novo* autoinflammatory manifestations were observed in 2 patients in the first months of FU.

### PLATELETS

Median platelet count increased from 18.0 (95% CI: 11.5-27.0)×10^9^/L at baseline (n=22), to 59.0 (95% CI: 39.0–84.0) ×10^9^/L at year 1 (n=23), and to 68.3 (95% CI: 60.5-112.0) ×10^9^/L at year 5 (n=22), with a sustained increase compared to baseline at all available later timepoints, up to year 13 (Fig. 4A). Sixteen (84.2%) out of 19 evaluable participants showed an improved platelet count at year 1. At all subsequent timepoints a sustained improvement was reported for at least 88% of evaluable participants (Table S15 and S16). Mean platelet volume (MPV) increased relative to baseline, with normal value in >90% of participants at year 1 and year 5. The annualized rate of platelet infusions decreased over time (Table S17) and sustained platelet transfusions were discontinued at a median of 46.5 days (range: 9-261 days) after treatment (Fig. 4B). An exploratory analysis of platelet activation profile indicated the restoration of platelet function after etu-cel infusion^14^.

**Figure 4.**
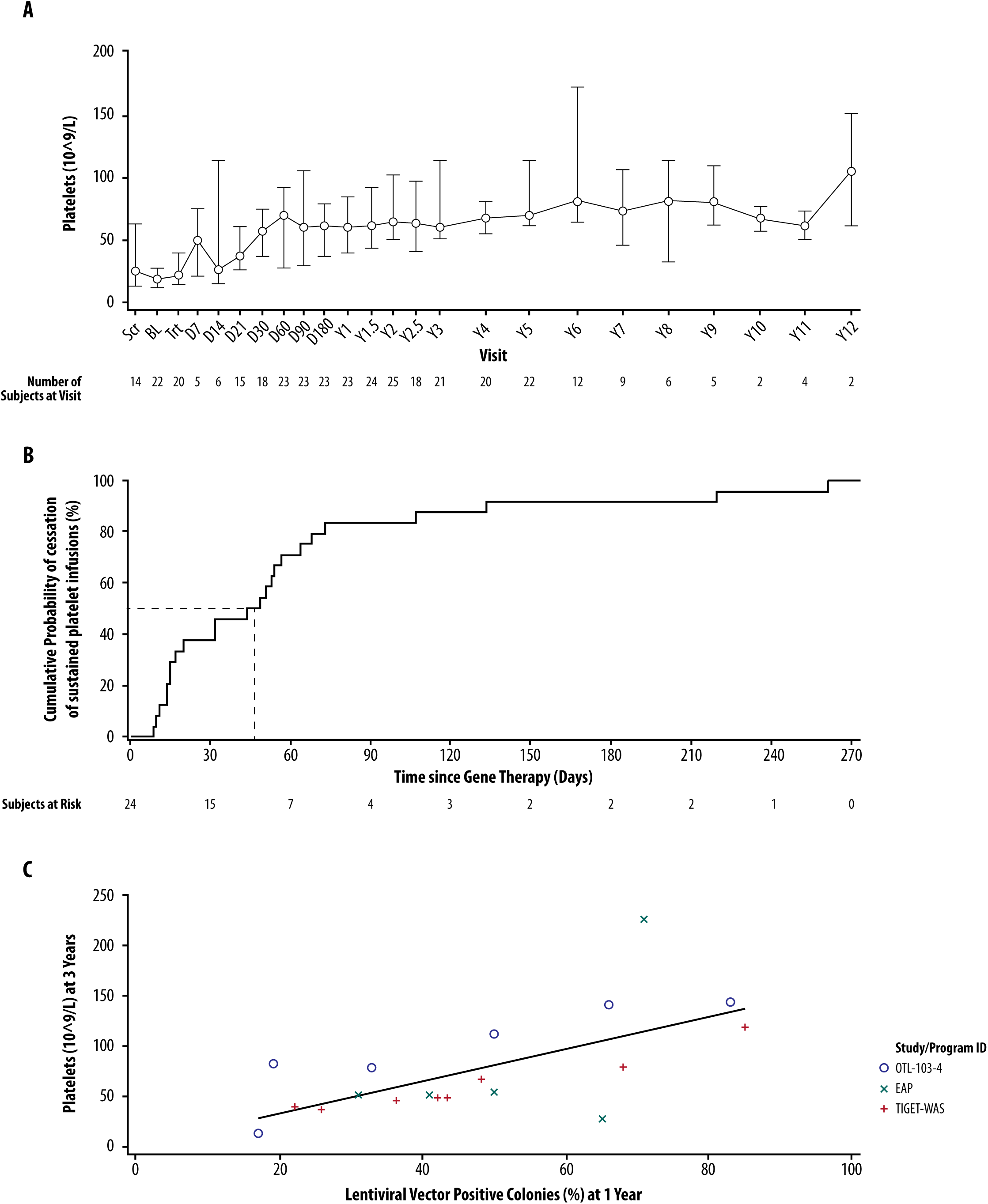
Improvement of thrombocytopenia after treatment with etu-cel (Efficacy population) **(A) Median (95% CI) values of platelet count over time.** Platelet samples taken within 7 days of a platelet transfusion, 25 days of romiplostim, or 14 days of eltrombopag were excluded from the calculation of data displayed in this figure. Results based on fewer than 2 participants were omitted. Note: BL=baseline; CI=confidence interval; D=day; Scr=Screening; Trt=Treatment (etu-cel); Y=Year. Transduced cells infusion (gene therapy) was performed at the Treatment Visit (Trt). **(B) Kaplan-Meier plot of time to cessation of sustained platelet infusions (Efficacy population)** Only participants who received platelet infusions post-etu cel were included in the analysis. The dashed line represents the median time of cessation of sustained platelet infusions. **(C) Correlation between platelet count at 3 years after etu-cel infusion and level of engraftment of gene corrected hematopoietic stem and progenitor cells in the BM at 1 year follow-up**, expressed as percentage (%) of lentiviral vector positive colonies. Note: BM = Bone Marrow, mPB = Mobilized Peripheral Blood, VCN = Vector Copy Number.

An exploratory analysis on covariates of efficacy showed a strong positive correlation between the years 2 and 3 platelet count and the percentage of LV-positive colonies at 1 year follow-up and between the year 2 platelet count and engraftment (BM CD34^+^ VCN/cell) at 1 year follow-up (Fig. 4C, Fig. S10). A positive correlation was also observed between the % of PB WASP+ platelets at year 2 and engraftment of LV+ colonies (Fig. S10C).

### SAFETY

Etu-cel was well tolerated. No treatment-related AEs were reported and there was no evidence of RCL, antibody response to WASP, or ACP. Insertion site analyses^15^ revealed polyclonal engraftment, with no signs of insertional mutagenesis, enrichment for proto-oncogenes or contraction of diversity (see Supplementary Results). Overall, AEs following treatment with etu-cel were consistent with those expected in WAS patients undergoing hematological reconstitution after RIC and not indicative for specific risk related to the DP. Fifty-seven serious adverse events (SAEs) occurred in 19 subjects in post-treatment phase (Table S18). Most SAEs (60%) occurred in the first 6 months post-GT and 54% of SAEs were infections, with most frequently device-related infections. One death occurred in an adult patient due to deterioration of a pre-existing form of iron accumulation encephalopathy, which was not considered related to etu-cel; a relationship with the conditioning regimen could not be ruled out by the treating physician (see Supplementary Results). One event of thyroid papillary carcinoma occurred in a patient almost 5 years after gene therapy, and was demonstrated to be not related to etu-cel. Relationship to conditioning could not be excluded since it is recognized that thyroid tumors can occur after chemotherapy for HSCT (see Supplementary Results). The most common AE ≥ grade 3 was device-related infection (Table S19 and S20). None of the treated patients required any secondary procedures (e.g. splenectomy, HSCT) after GT.

## DISCUSSION

The long-term follow-up data from patients treated with a single infusion of etu-cel provides compelling evidence that this LV HSC GT offers durable marked clinical benefit and an excellent safety profile in WAS patients. Our study represents the largest WAS population treated with GT with the longest follow-up available to date (up to 13 years with a median of 5.7 years).

A major clinical hallmark of WAS is the occurrence of recurrent, often severe, infections due to the underlying immune deficiency. The integrated analysis shows a profound reduction in the incidence of severe infections after GT, with protection maintained until last observation in all patients. This improvement reflects the successful engraftment of corrected progenitors and the restoration of immune function. Patients progressively achieved immune reconstitution, discontinued immunoglobulin replacement therapy, responded to vaccination mounting protective antibody responses against pathogens and vaccines, in line with the correction of B-cell defects after gene therapy^16–18^.

The restoration of normal immune functions translated into multiple clinical benefits, with resolution of eczema and autoimmunity in most patients, progressive improvement of social life and reduced morbidity. Transient manifestations of autoimmune cytopenia were observed shortly after GT in 18.5% of patients, while 14.8% of patients showed autoimmune manifestations > 6 months after etu-cel infusion, which resolved after short-term treatment or without intervention. Autoimmune events have been reported in 14% to 20% of patients with WAS after HSCT^7,16,19^ and were also observed in 2 out of 5 patients post-GT in a distinct phase I/II study which used a similar vector backbone but did not include rituximab as preparatory conditioning^18^. A delayed reconstitution of T and/or B regulatory cells has been suggested to contribute to slower or incomplete correction of immune dysregulation after GT or HSCT^18^.

Thrombocytopenia and bleeding events are pathognomonic of all forms of WAS^1,4,5^. Our results show that after etu-cel infusion nearly all patients experienced a sustained increase in platelet count, normalization of platelet volume and functional assays indicated recovery of platelet activity^14^, with a substantial and sustained reduction of severe and moderated bleedings. Importantly, in 96% of patients, platelet count at last follow-up was ≥ 50 ×10^9^/L, allowing omission of activity restrictions to avoid trauma-associated bleeding and prophylactic treatment in case of surgery^20,21^. Exploratory analyses suggested that a higher level of engraftment of gene corrected progenitors positively influences the production of normal platelets by transduced megakaryocytic lineage. This is in line with data from other studies which showed a correlation between platelet counts and gene marking in myeloid cells or BM.

The improvement of immune function, reduction of bleeding risk and cessation of sustained supportive treatments allowed the patients to normalize their social lives.

The safety profile of etu-cel is reassuring. Adverse events were largely attributable to conditioning or device-related infections and were manageable in the clinical context. Unlike earlier trials performed with γ-retroviral vectors to treat WAS^22,23^, the use of self-inactivating lentiviral vectors and the choice of a promoter reconstituted from the endogenous WAS locus to drive WASp expression was not associated with insertional oncogenesis nor clonal expansion. Comprehensive integration site analyses consistently confirmed polyclonal hematopoiesis. Our data reinforce the recommendation of using a moderately active, physiological promoter in the LV backbone, as adopted in several other HSC gene therapy trials^23–25^ and approved products^26^, which have reported similar safety as shown here for WAS, rather than strong retroviral promoters, as used for the treatment of X-linked adrenoleukodystrophy (ALD), where the occurrence of a significant rate of insertional oncogenesis has been reported^27–29^.

Importantly, the efficacy and safety outcomes were consistent across the heterogeneous study population, demonstrating similar results irrespective of age at the time of treatment, baseline disease characteristics, underlying WAS mutations, or drug product formulation. In particular, the use of a cryopreserved formulation did not affect the outcome while it facilitates distribution, quality testing and timing of administration of etu-cel. Limitations of our study include lack of blinding and absence of a control group of patients treated with current standard of care. These limitations are inevitable due to ethical and feasibility issues, and justifiable for trials of GT in rare diseases^30^.

Recent reports from multicenter observational studies from international registries show overall survival after HSCT of about 90% with the majority of recipients achieving high-level donor chimerism and normal platelet counts^7,8^. Limitations due to donor availability, risk of rejection and GvHD, and higher morbidity in older patients remain a concern for HSCT. Results of our analyses suggest that etu-cel compares favorably with allogeneic HSCT, with absence of GvHD and graft rejection and no need of secondary procedures. While HSCT remains an established curative option, etu-cel could be considered for all WAS patients lacking a related HLA-identical donor.

A distinctive aspect of this program is that it originated in an academic center, was further developed by pharmaceutical companies and eventually licensed back to Fondazione Telethon under a not-for-profit model^31^. This framework underscores the potential to deliver innovative therapies for ultra-rare diseases, ensuring that patients benefit from cutting-edge science while minimizing barriers related to commercial prioritization^32^.

In conclusion, the observed immune and hematological improvements establish GT as a comprehensive disease-modifying intervention. The robust and reproducible effect supports etu-cel as a therapeutic option for a broad spectrum of WAS patients.

## Funding Statement

The clinical studies in the etu-cel development program are sponsored by Fondazione Telethon. The integrated analysis was conducted through a collaboration between the sponsor and the investigators. Study TIGET-WAS was sponsored until January 2013 by Fondazione Centro San Raffaele del Monte Tabor, from 2013 to 2016 by IRCCS San Raffaele Scientific Institute, by GlaxoSmithKline (GSK) from April 2016 to August 2018, and from 2018 to September 2023 by Orchard Therapeutics. Study OTL-103-4 was sponsored by Orchard Therapeutics until October 2023. Funding for the EAP (HE and CUP) was provided by Fondazione Telethon, GSK, and Orchard Therapeutics.

AA is the recipient of the Kröner Fresenius Prize for Medical Research 2020.

## Data Availability

Because of the small number of participants in the
study and potential for identification, individual
patient data beyond what is included in the
manuscript will not be available

## Acknowledgements

The authors would like to thank Prof. Maria Grazia Roncarolo, who started the program and was the initial clinical principal investigator of Study TIGET-WAS, Loïc Dupre and Samantha Scaramuzza who contributed to the preclinical development, and the employees of GSK and Orchard Therapeutics who participated to the clinical development program.

We wish to acknowledge the Telethon Foundation for continuous support and strategic guidance. We thank the clinical research nurses; the medical and nursing team of the Pediatric Immunohematology Unit, Stem Cell Transplant Program, Therapeutic Apheresis Unit and Cell Manipulation Lab, and Pediatric Unit of the IRCCS San Raffaele Scientific Institute, and of the Children’s Hospital of Atlanta (CHOA) for their professional care of patients during hospitalisation; the team of the Department of Anesthesia and Neurointensive Care for support; Margherita Levi, Samih El Hossary, cultural mediators, and all personnel and volunteers of Fondazione Telethon “Just like home” Program for constant support with families and loving care of the children; Mrs Giuliana Tomaselli, Luisella Meroni and Francesca Cancelli for administrative assistance; the staff of the SR-Tiget Clinical Trial Office for its support in trial management; the San Raffaele quality team, the lab staff of the SR-Tiget Clinical Laboratory; and the team of AGC Biologics SpA for manufacturing the lentiviral vector and drug product for etu-cel. We thank the physicians from 16 countries worldwide who referred patients and continued their follow-up with great commitment. This study is dedicated to all the patients with WAS and their families, whose unwavering support and trust has sustained the WAS GT program over the past 20 years.

